# Bushfire-smoke trigger hospital admissions with cerebrovascular diseases: evidence from 2019-20 bushfire in Australia

**DOI:** 10.1101/2023.04.18.23288775

**Authors:** Md Golam Hasnain, Carlos Garcia-Esperon, Yumi Tomari, Rhonda Walker, Tarunpreet Saluja, Md Mijanur Rahman, Andrew Boyle, Christopher R Levi, Ravi Naidu, Gabriel Filippelli, Neil J Spratt

**Author notes:** **Corresponding author** Md Golam Hasnain, School of Medicine and Public Health, College of Health, Medicine, and Wellbeing, University of Newcastle, University Drive, Callaghan, NSW 2308, Australia.

## Abstract

**Background:** Exposure to ambient air pollution is strongly associated with increased cerebrovascular diseases. The 2019–20 bushfire season in Australia burnt 5.4 million hectares of land in New South Wales alone, with smoke so severe it affected cities in Argentina, 11,000 km away. We aimed to determine the effects of i) short-term air pollution triggered by bushfires and ii) high smoke days in increasing the daily number of hospital admissions with cerebrovascular diseases.

**Methods:** Hospitalisation data were accessed from the admitted patient dataset from seven local Government areas of Hunter New England Local Health District. The bushfire period was defined from 1 October 2019 to 10 February 2020, and a similar period from 2018-19 as the control. High bushfire smoke days were days when the average daily concentration of particulate matter was higher than the 95^th^ percentile of the control period. Poisson regression models and fixed effect meta-analysis were used to analyse the data.

**Results:** In total, 275 patients with cerebrovascular admissions were identified, with 147 (53.5%) during the bushfire (2019-20) and 128 (46.5%) in the control period (2018-19). There was no significant increase in daily cerebrovascular disease (Incidence Rate Ratio, IRR: 1.04; 95% CI: 0.98-1.05; p-value: 0.73) or ischemic stroke (IRR: 1.18; 95% CI: 0.87-1.59; p-value: 0.28) admissions over the entire bushfire period. However, the high bushfire smoke days were associated with increased ischaemic stroke-related hospital admissions with a lag of 0-1 days (IRR: 1.28; 95% CI: 1.01-1.62; I^2^=18.9%). In addition, during the bushfire period, particulate matter, both PM_10_ and PM_2.5_ (defined as particulates that have an effective aerodynamic diameter of 10 microns, and 2.5 microns, respectively), were also associated with increased ischaemic stroke admissions with a lag of 0 to 3 days.

**Conclusion:** The results suggested an association between particulate matter and high smoke days with increased hospital admissions due to cerebrovascular diseases during the recent Australian bushfire season.

## Introduction

Bushfire is an inevitable and ever-increasing challenge in many parts of the world.^1-2^ Global warming increases the number of extremely hot and dry days in some regions, leading to more frequent episodes of bushfires.^3^ Australia suffered an extensive bushfire season in 2019-20. From October 2019 to March 2020, fires burnt an estimated 18.6 million hectares across Australia, with New South Wales being the most affected state (5.4 hectares of land).^4-5^

Bushfire smoke contains both gaseous and particulate matter.^6^ The effect of particulate matter, including total inhalable particulate matter (PM_10_) and the finer particulate matter fraction (PM_2.5_), defined as particulates have an effective aerodynamic diameter of 10 microns, and 2.5 microns, respectively, are of great concern because of their presence in high concentrations in smoke and their potential impact on health.^7^ Fire days may increase mortality and morbidity, premature death, and exacerbation of cerebrovascular conditions.^8^ Moreover, there is significant evidence from other settings of an association between higher particulate matter and ischaemic stroke incidence. Exposure to particulate matter, PM_10_ and/or PM_2.5_ is associated with an increased risk of stroke-related hospitalisations and death.^9^ Another study evaluating the 2019-20 bushfire effect on eastern Australia estimated that bushfire smoke accounted for 1124 (95% CI, 211–2047) emergency hospitalisations related to cardiovascular conditions, including strokes and other cerebrovascular diseases.^10^ But there was insufficient information about how patients with a confirmed cerebrovascular condition responded to bushfire smoke, and what the related health impacts of PM_10_ and PM_2.5_ were during the bushfire season and smoke days in 2019 and 2020. Therefore, this study aims to

- examine the association between particulate matter concentrations and cerebrovascular disease hospital admissions during the 2019-20 bushfire period with the previous similar period in 2018-19 (Aim 1),
- compare the number of hospitalisations following high bushfire smoke days with those following non-smoke days (Aim 2), and
- study the association between particulate matter concentrations and cerebrovascular disease hospital admissions during the 2019-20 bushfire period (Aim 3).

## Methods

### Study area and population

The study was conducted in Hunter New England Local Health District (HNE-LHD), New South Wales (NSW), Australia, covering an area of 131,785 km^2^ and 920,370 people. It encompasses a metropolitan centre (Newcastle) surrounded by a large, low-density rural area with some small cities or towns.^11^ This study focused on seven Local Government Areas (LGAs) within the health district: Newcastle, Muswellbrook, Singleton, Tamworth, Armidale, Narrabri, and Gunnedah (Figure 1). These areas were chosen because they have at least one on-site air pollution monitoring measuring PM_10_ and PM_2.5_. These seven LGAs had a population of 318,323 people (Newcastle LGA 160,919, Tamworth LGA 60,998, Armidale 30,311, Singleton 23,595, Muswellbrook 16,468, Narrabri 13,481, and Gunnedah 12,551 people), representing 35% of the population of the HNE-LHD.^12^

**Figure 1:**
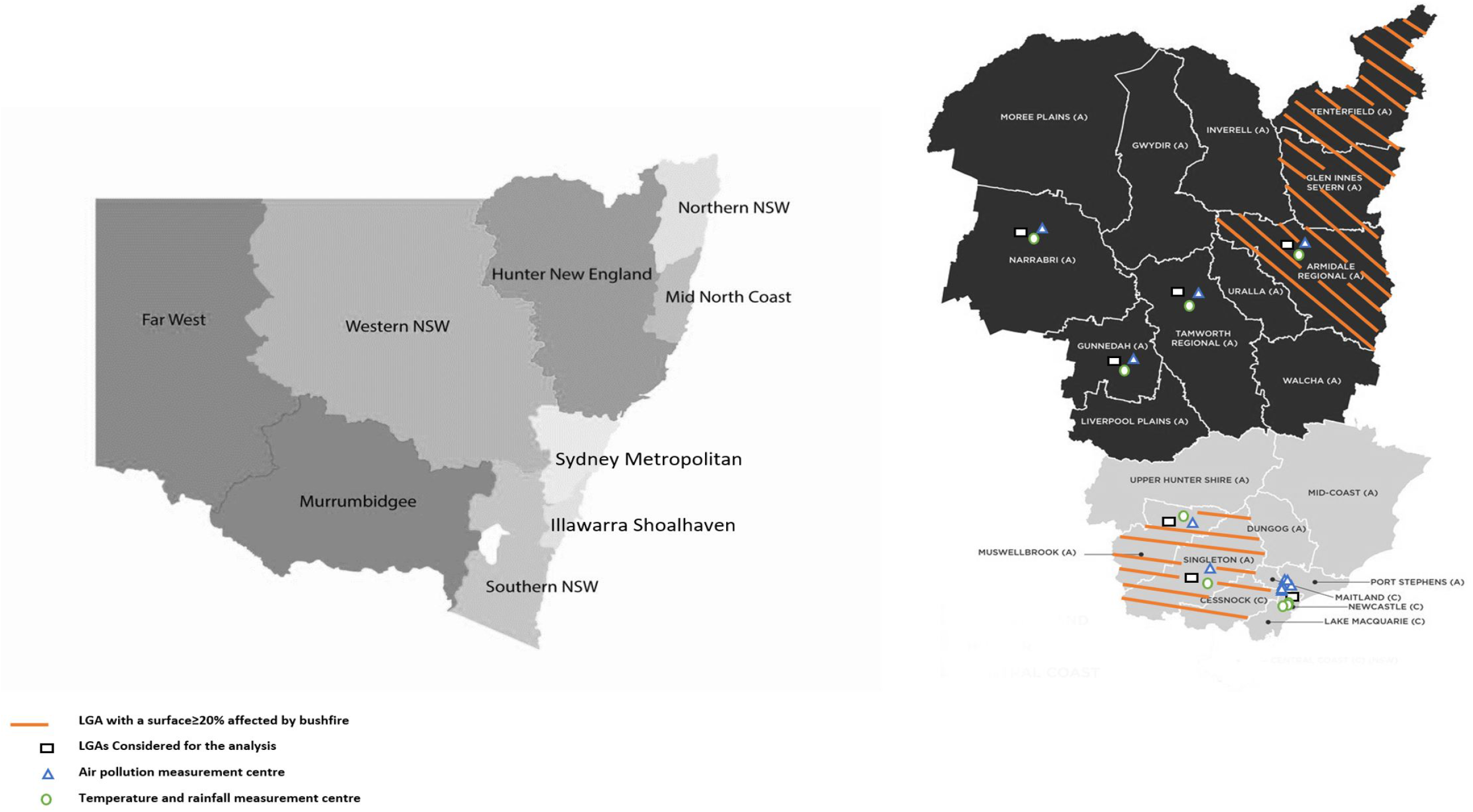
Diagram showing the distribution of bushfire 2019-2020 affected areas and weather measurement centres across HNE-LHD.^16-18^

### Bushfire seasons and smoke days

The 2019-2020 bushfire season started from October 2019 to February 2020. These affected multiple regions in Australia, especially within NSW and in the Hunter region. Most of the fires were extinguished by 10 February 2020.^10^ Therefore, we defined the bushfire period as extending from 1 October 2019 to 10 February 2020. The period from 1 October 2018 to 10 February 2019 was designated the control period.

Smoke exposure during the bushfire varies enormously, depending on the proximity of fires and prevailing wind and weather conditions. To determine the effects of bushfire smoke, we defined high bushfire smoke and non-smoke days as follows:

- <u>High bushfire smoke days</u>: Days from the bushfire period when the daily concentration of either PM_10_ or PM_2.5_ was greater than the 95^th^ percentile of the control period. Population-weighted control period pollutant data was used to calculate the percentiles.
- <u>Bushfire non-smoke days</u>: All the days from the control period were considered non-smoke days.

### Hospitalisation and Patient data

Hospitalisation with cerebrovascular disease was the primary outcome variable. Hospitalisation data were retrieved from the HNE-LHD local electronic database for admitted patients, which includes information on discharges from all publicly funded hospitals within the LHD. All cases were coded according to the World Health Organization’s (WHO’s) International Classification of Diseases, 10th Revision (ICD-10).^13^ Cerebrovascular diseases were coded as I60-I69 and ischemic stroke as I63. Daily counts of patients with an address within the seven selected LGAs with a primary diagnosis identified on discharge corresponding to one of the ICD-10 codes of interest were obtained. The date of admission was used as the incidence date for analysis, and this date was considered the day of pollutant exposure which was also used as an index day to calculate lag and lead days.

### Air pollution and meteorological data

Daily mean concentrations of air pollutants data (PM_10_ and PM_2.5_) were collected from the NSW Government Planning, Industry and Environment website^14^, for 12 measuring stations within the five selected LGAs: Newcastle, Beresfield, Wallsend, Carrington, Mayfield, Stockton, Muswellbrook, Singleton, Tamworth, Armidale, Narrabri, and Gunnedah. The daily maximum temperature and daily total rainfall data were collected from the Australian Government Bureau of Meteorology website for nine weather stations within the same LGAs (Figure 1).^15^

### Ethics Approval

Ethics approval was taken from the Hunter New England Health Human Research Ethics Committee (2020/ETH01801).

### Statistical analysis

Initial analysis was performed using descriptive statistics, with frequency and percentages for categorical variables and mean and standard deviation (SD) or median and interquartile range (IQR) for continuous variables. The patient characteristics between bushfire and control were compared using Chi-square tests (frequency)/t-test (mean)/Kruskal-Wallis test (median). Poisson regression or negative binomial regression models were used where appropriate to measure the effect of bushfire period, smoke days and particulate matters on cerebrovascular hospitalisation. Each regression model was adjusted for daily maximum temperature and daily total rainfall. Separate models were performed for each outcome. Considering the possible delayed and or cumulative effects of air pollution, we created a cross-basis matrix for air pollutants within the distributed lag linear model framework. The selection of lag days was justified by the information criteria: Schwarz’s Bayesian information criterion (SBIC), Akaike’s information criterion (AIC), and the Hannan and Quinn information criterion (HQIC). The lag length was initially set as ten days; the results showed that the lag effect mainly lasted for three days (S1). Finally, we considered three days as a lag length in the later analysis to measure the effect of PM_10_ and PM_2.5_ on the daily number of hospital admission during the bushfire period. The effect of high bushfire smoke days was calculated for each LGAs separately. The outcome variables were the cumulative number of daily hospital admission at a lead of 0–3 days, which admission day was considered as “Day 0”. A fixed effect meta-analysis was conducted to calculate the overall effect of high bushfire smoke days. Robust standard errors were used to control for mild violations under assumptions. Results are presented as an incidence rate ratio (IRR) and 95% confidence intervals (95% CI). A p-value < 0.05 was considered significant. All analyses were performed with Stata Statistical Software, version 15 (Stata Corp, College Station, TX, USA).

## Results

In total, 275 patients with cerebrovascular disease admissions were identified during the bushfire and control period (128, 46.5% in the control period and 147, 53.5% in the bushfire period). Of those, 182 (66.2%) were ischemic strokes, 97 (53.3%) occurred during the bushfire period and 85 (46.7%) during the control period. Furthermore, 12 (4%) were diagnosed as subarachnoid haemorrhages, 34 (12%) as intracerebral haemorrhages, 17 (6%) as subdural haemorrhages and 30 (11%) were codified as stroke (unspecified subtype).

The median age of patients with a cerebrovascular disease admission was 75 [65-84] years, and 154 (56%) were female. There were no major differences in age, sex, or demographic variables, between the bushfire and control period patients (Table 1). In the subgroup of patients with an ischemic stroke, the median age was 74.5 [66-85] years, and 101 (55.5%) were female. There were no differences between the baseline characteristics of these patients from the bushfire period and those from the control period. In addition, substantial differences in the daily distribution of particulate matter were observed between the bushfire and control periods, S2.

**Table 1:** Description of patient characteristics across groups of cerebrovascular diseases.

|  |  | Cerebrovascular Diseases |  |  | Ischemic Stroke |  |  |
| --- | --- | --- | --- | --- | --- | --- | --- |
|  |  | N = 275 |  |  | N = 182 |  |  |
|  |  | *Bushfire* | *Control* | *p-value* | *Bushfire* | *Control* | *p-value* |
|  |  | *N = 147* | *N = 128* |  | *N = 97* | *N = 85* |  |
| **Age in years** |  |  |  |  |  |  |  |
| - | Median (Q<sub>1</sub>-Q<sub>3</sub>) | 75 (66-84) | 75 (64.5-85) | 0.917 | 73 (66-83) | 76 (65-86) | 0.38 |
| **Sex** |  |  |  |  |  |  |  |
| - | Female, n (%) | 78 (53) | 76 (59) | 0.293 | 50 (51) | 51 (60) | 0.25 |
| **Living with partner** |  |  |  |  |  |  |  |
| - | Yes, n (%) | 74 (50) | 63 (49) | 0.463 | 51 (53) | 42 (49) | 0.81 |
| **Identified as Aboriginal** |  |  |  |  |  |  |  |
| - | Yes, n (%) | 6 (4) | 9 (7) | 0.283 | 2 (2) | 6 (7) | 0.10 |
| **Speaking language** |  |  |  |  |  |  |  |
| - | English, n (%) | 144 (98) | 123 (96) | 0.641 | 95 (98) | 81 (95) | 0.30 |

\*p-value <0.05 considered significant

### Effect of bushfire period

There were no significant changes in total daily cerebrovascular diseases admissions (IRR: 1.04; 95% CI: 0.98-1.05; p-value: 0.73) or ischemic stroke admissions (IRR: 1.18; 95% CI: 0.87-1.59; p-value: 0.28) over the entire bushfire period Figure 2.

**Figure 2:**
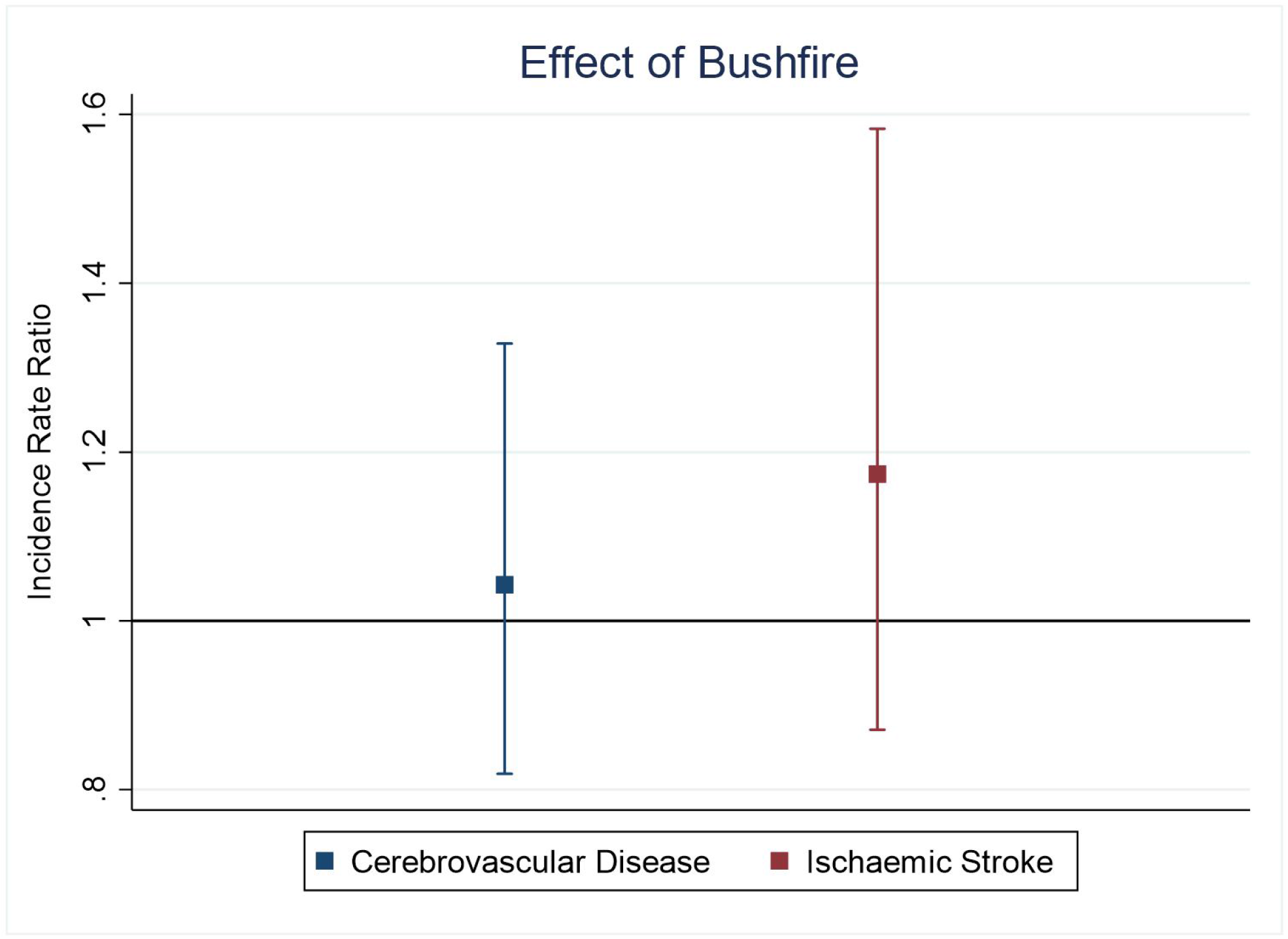
Effect of bushfire period on cerebrovascular diseases and ischemic stroke.

### Effect of bushfire high smoke days

The meta-analysis showed that high bushfire smoke days were associated with a 28% increase in the cumulative daily number of ischaemic stroke-related hospital admissions across a lead of 0-1 days (IRR: 1.28; 95% CI: 1.01-1.62; I^2^=18.9%); Figure 3. A similar but non-significant trend was also observed for cerebrovascular disease-related hospital admissions, S3. In addition, stratified findings showed a similar positive trend for most of the included LGAs except Singleton and Narrabri.

**Figure 3:**
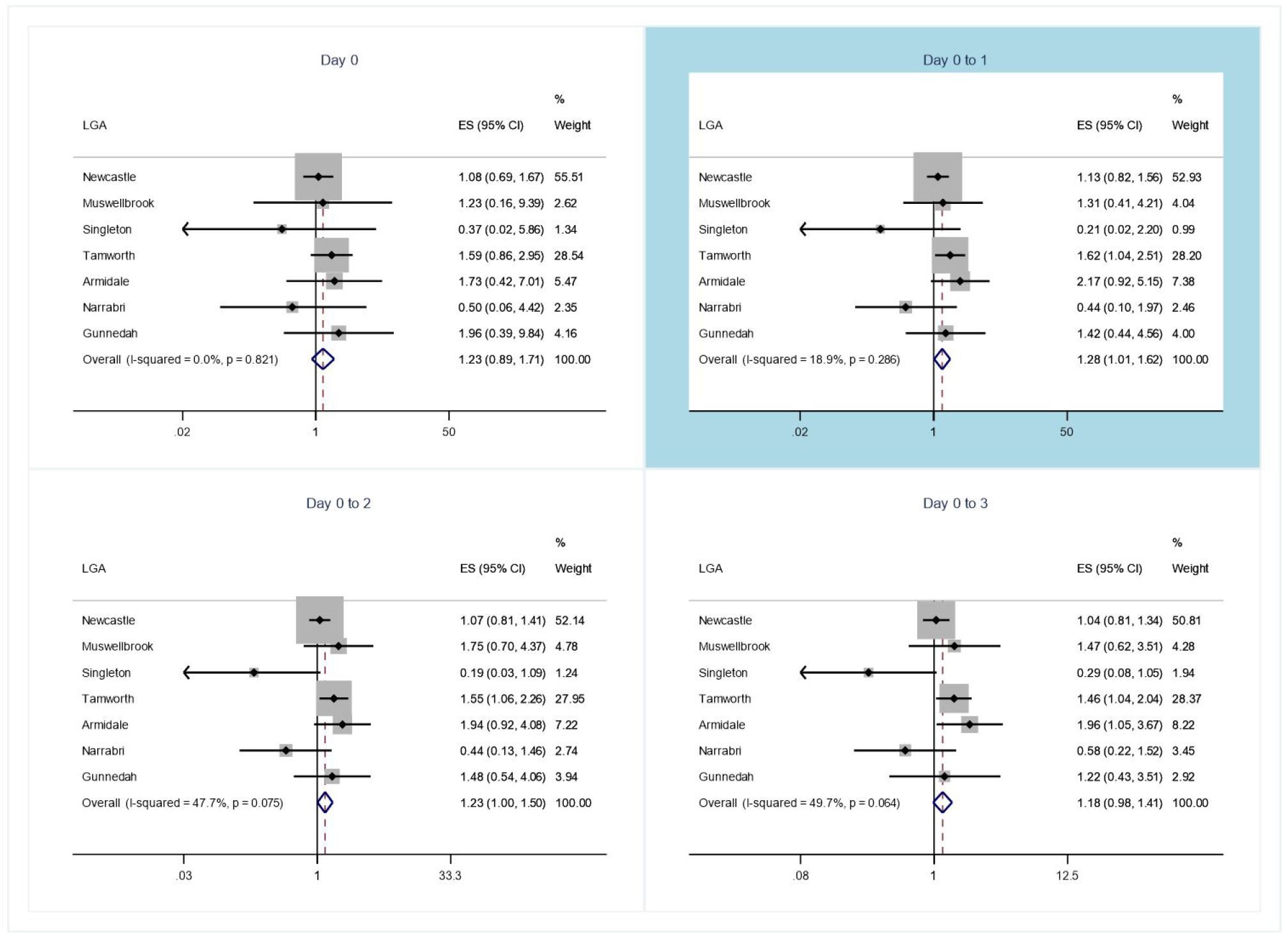
Effect of high bushfire smoke days on a cumulative number of daily hospital admissions due to ischaemic stroke across a lead of 0 to 3 days.

### Effect of PM_10_ and PM_2.5_ during bushfire and control periods

During the bushfire period, PM_10_ was associated with increased ischaemic stroke admissions across a lag of 0 to 3 days, Table 2. The effect of PM_2.5_ was only found significant for the lag 0-2 (IRR: 1.01; 95% CI: 1.00-1.03; p-value: 0.03). No significant effect was observed during the control period between PM_10_ or PM_2.5_ and the number of total daily admissions of cerebrovascular disease or ischemic stroke; Table 2.

**Table 2:**
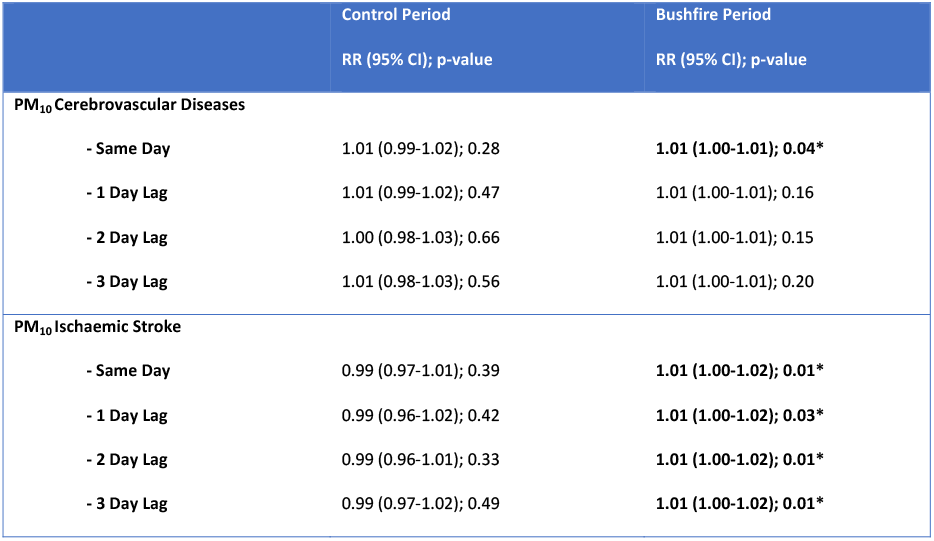
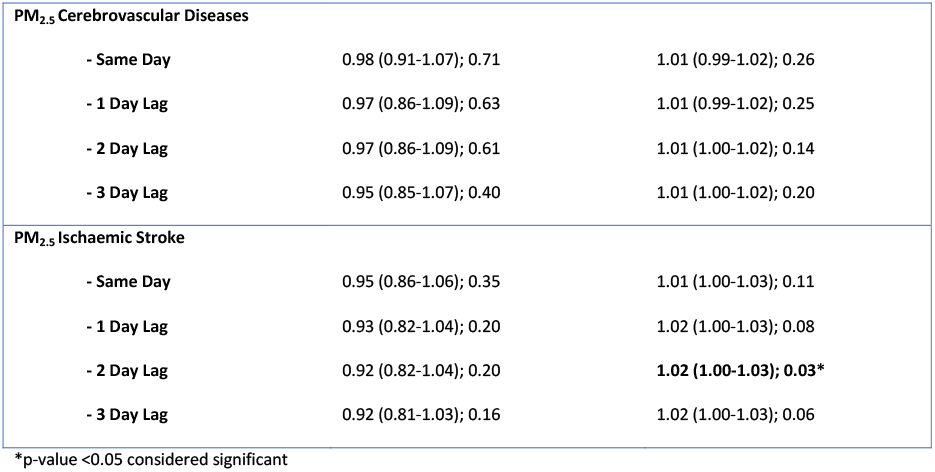
Effect of PM_10_ and PM_2.5_ on daily hospital admissions due to cerebrovascular diseases and ischemic stroke across lag 0-3 days.

|  |  | Control Period | Bushfire Period |
| --- | --- | --- | --- |
|  |  | RR (95% CI); p-value | RR (95% CI); p-value |
| **PM<sub>10</sub> Cerebrovascular Diseases** |  |  |  |
| - | **Same Day** | 1.01 (0.99-1.02); 0.28 | **1.01 (1.00-1.01); 0.04\*** |
| - | **1 Day Lag** | 1.01 (0.99-1.02); 0.47 | 1.01 (1.00-1.01); 0.16 |
| - | **2 Day Lag** | 1.00 (0.98-1.03); 0.66 | 1.01 (1.00-1.01); 0.15 |
| - | **3 Day Lag** | 1.01 (0.98-1.03); 0.56 | 1.01 (1.00-1.01); 0.20 |
| **PM<sub>10</sub> Ischaemic Stroke** |  |  |  |
| - | **Same Day** | 0.99 (0.97-1.01); 0.39 | **1.01 (1.00-1.02); 0.01\*** |
| - | **1 Day Lag** | 0.99 (0.96-1.02); 0.42 | **1.01 (1.00-1.02); 0.03\*** |
| - | **2 Day Lag** | 0.99 (0.96-1.01); 0.33 | **1.01 (1.00-1.02); 0.01\*** |
| - | **3 Day Lag** | 0.99 (0.97-1.02); 0.49 | **1.01 (1.00-1.02); 0.01\*** |

|  |  |  |
| --- | --- | --- |
| **PM<sub>2.5</sub> Cerebrovascular Diseases** |  |  |
| - Same Day | 0.98 (0.91-1.07); 0.71 | 1.01 (0.99-1.02); 0.26 |
| - 1 Day Lag | 0.97 (0.86-1.09); 0.63 | 1.01 (0.99-1.02); 0.25 |
| - 2 Day Lag | 0.97 (0.86-1.09); 0.61 | 1.01 (1.00-1.02); 0.14 |
| - 3 Day Lag | 0.95 (0.85-1.07); 0.40 | 1.01 (1.00-1.02); 0.20 |
| **PM<sub>2.5</sub> Ischaemic Stroke** |  |  |
| - Same Day | 0.95 (0.86-1.06); 0.35 | 1.01 (1.00-1.03); 0.11 |
| - 1 Day Lag | 0.93 (0.82-1.04); 0.20 | 1.02 (1.00-1.03); 0.08 |
| - 2 Day Lag | 0.92 (0.82-1.04); 0.20 | **1.02 (1.00-1.03); 0.03\*** |
| - 3 Day Lag | 0.92 (0.81-1.03); 0.16 | 1.02 (1.00-1.03); 0.06 |

\*p-value <0.05 considered significant

## Discussion

The research found no correlation between the overall bushfire and the control period and an increase in the daily number of hospital admissions for ischemic stroke or cerebrovascular diseases. However, compared to non-smoke days, a substantial rise in hospital admissions was seen during days with high bushfire smoke. The study also showed that particulate matter (PM_10_ and PM_2.5_ separately) significantly increased the daily number of hospital admissions during the bushfire period, while no such effect was seen during the control period.

When the bushfire period was compared to the control period, the study found no significant difference. According to the Australian Institute of Health and Welfare (AIHW), these findings are in line with what was found.^4^ After analysing data from the 2019-2020 bushfire season, the AIHW found that there was a small increase in the number of patients presenting with symptoms of cerebrovascular illness at NSW emergency departments. There was no discernible pattern to the growth, though. This study, however, found that days with high bushfire smoke were linked to more hospitalizations than days with non-smoke days. There has been a dearth of such findings in recent publications related to 2019-20 bushfire.^4,10^ We think this additional information uncovers a substantial concern with the data that has been gathered. It is extremely challenging to link specific exposures to subsequent hospitalisations. By its very nature, bushfire smoke exposure is highly variable from one location to the next; individual exposure is also highly variable since remaining indoors, for example, greatly reduces exposure. Therefore, the finding of a significant effect on the worst bushfire days, when using only limited numbers of monitoring stations, suggests that this may be quite a substantial effect.

The study also identified a significant effect of particulate matter, both PM_10_ and PM_2.5,_ in increasing the number of hospitalisations during the bushfire period. Particulate matters are known to significantly increase the number of strokes and cerebrovascular disease-related hospital admissions.^19^ Exposure to particulate matter increases inflammation, antioxidant activity and circulating blood platelet activation and decreases vascular endothelial functions and enzyme activity, which may increase peripheral thrombosis and blood clotting.^20-22^ Over the course of ten years or more, two investigations from Australia looked at whether or not exposure to particulate matter during bushfire smoke days increased hospital admissions for cerebrovascular disorders or stroke.^23-24^ Hospitalizations due to cerebrovascular or stroke-related conditions were not linked to the higher concentration of particulate matter, according to the investigations. Previous research had mixed results because of methodological differences and outcome measurement; for example, they counted transient ischaemic attack patients as having a stroke and characterised bushfire smoke days using the 99^th^ percentile of the entire study period. Patients with a confirmed diagnosis were included in our study, and smoking days were defined using data from the control group alone. Furthermore, the most recent devastating bushfire in 2019-2020 occurred almost a decade after the prior research were completed. Consequently, a shift in demographic and environmental factors may also account for this discrepancy. Finally, although this study demonstrated increased particulate matter concentrations during the bushfire period, some days from the control period (three days) also showed high concentrations of PM_10_. Those days might be associated with some locally sparked bushfires within the Hunter region (Kurri Kurri and Port Stephens) controlled quickly, within days, by firefighters.^25^ This study’s primary strength is its evaluation of the time lag between exposure and effect during a bushfire and its estimation of the number of admissions based on confirmed diagnoses. More crucially, the study took place in an area with a somewhat consistent population and habitat, making it possible to determine an effect size more accurately. Nevertheless, the study has several limitations. Assessing the consequence of fire smoke exposure on health is always a big challenge. The sporadic characteristics of significant fire events make it challenging to distinguish when and where air quality will be poor accurately. Many affected areas do not have routine air quality monitoring stations. In addition, the peak exposures may be too short-lived to detect all but the most sensitive health outcomes with adequate statistical power. Because of these local effects and the effect of variable time spent outdoors, some degree of exposure misclassification is inevitable, which would likely have biased our results towards the null. Another potential limitation, as with most studies of this type, is fixed-site monitors. It is possible that available monitors did not detect smaller smoke plumes on some occasions and did not take into account geomorphic and atmospheric characteristics that tend to dilute, or concentrate, particulate matter in certain areas, and these would not have been classified as severe smoke events. Finally, the models used to generate results were single pollutant models and not adjusted for other pollutants, which needs to be considered for future large-scale studies. Future research should consider evaluating therapies designed to lower risk, especially for those with a higher risk of stroke. For example, reducing the impact of particulate matters by limiting outdoor activities, utilising an air purifier with a high-efficiency particle air filter, and wearing masks (P2 or N95 masks with caution).

This study found that exposure to high concentrations of particulate matter during intense bushfires and exposure to days with high levels of bushfire smoke were consistently associated with increased rates of cerebrovascular disorders and stroke-related admissions. As a result, during the season of significant bushfires, the required precautions should be taken to reduce the risk of being exposed to smoke and the severity of the disease load.

## Data Availability

The data that support the findings of this study are available on request from the corresponding author.

## Acknowledgments

None

## Sources of funding

None

## Disclosures

None

## Supplementary material

Table S1

Figure S2-S3

## Notes

### Competing Interest Statement

The authors have declared no competing interest.

### Clinical Trial

NA

